# Higher education responses to COVID-19 in the United States: Evidence for the impacts of university policy

**DOI:** 10.1101/2021.10.07.21264419

**Authors:** Brennan Klein, Nicholas Generous, Matteo Chinazzi, Zarana Bhadricha, Rishab Gunashekar, Preeti Kori, Bodian Li, Stefan McCabe, Jon Green, David Lazer, Christopher R. Marsicano, Samuel V. Scarpino, Alessandro Vespignani

## Abstract

With a dataset of testing and case counts from over 1,400 institutions of higher education (IHEs) in the United States, we analyze the number of infections and deaths from SARS-CoV-2 in the counties surrounding these IHEs during the Fall 2020 semester (August to December, 2020). We used a matching procedure designed to create groups of counties that are aligned along age, race, income, population, and urban/rural categories—socio-demographic variables that have been shown to be correlated with COVID-19 outcomes. We find that counties with IHEs that remained primarily online experienced fewer cases and deaths during the Fall 2020 semester; whereas before and after the semester, these two groups had almost identical COVID-19 incidence. Additionally, we see fewer deaths in counties with IHEs that reported conducting any on-campus testing compared to those that reported none. We complement the statistical analysis with a case study of IHEs in Massachusetts—a rich data state in our dataset—which further highlights the importance of IHE-affiliated testing for the broader community. The results in this work suggest that campus testing can itself be thought of as a mitigation policy and that allocating additional resources to IHEs to support efforts to regularly test students and staff would be beneficial to mitigating the spread of COVID-19 in the general population.

---

In the United States over 19.6 million people attend institutes of higher education (IHEs; i.e., colleges, universities, trade schools, etc.) [1], where students often live in highly clustered housing (e.g. dorms), attend in-person classes and events, and gather for parties, sporting events, and other large gatherings. Because of this, the COVID-19 pandemic presented a particular challenge for IHEs during the Fall 2020 semester [2–7]. On the one hand, bringing students back for on-campus and in-person education introduced the risk that an IHE would contribute to or exacerbate large regional outbreaks [8–16]; on the other hand, postponing students’ return to campus may bring economic or social hardship to the communities in which the IHEs are embedded [17–20], since IHEs are often large sources of employment for counties across the United States. As a result, IHEs instituted a variety of “reopening” strategies during the Fall 2020 semester [21–32]. Studying the different impacts of IHE policies on the surrounding community is made even more difficult because of the lack of a centralized data source and standardized reporting style. Among IHEs that brought students and employees back to campus—either primarily in person or in a “hybrid” manner—we see different approaches to regularly conducting (and reporting) COVID-19 diagnostic testing for students, faculty, and staff throughout the semester.

Here, we introduce a dataset of testing and case counts from over 1,400 IHEs in the United States (Figure 1), and we use this dataset to isolate and quantify the impact that various IHE-level policies may have on the surrounding communities during the Fall 2020 semester (August to December, 2020). The Campus COVID Dataset [33] (see Data & Methods section), collected manually through the COVID dashboards of over 1,400 IHEs, allows the comparison of COVID-19 outcomes between counties with IHEs where students returned to fully or *primarily in-person* education and counties with IHEs that remained fully or *primarily online*. IHE reopening status is based on data from [23]. We group the categories of “fully online” and “primarily online” into “primarily online” and do the same for fully/primarily in-person; schools listed as “hybrid” are not included in this comparison but present interesting avenues for future research. Second, we quantify the benefits of IHE-affiliated testing by comparing COVID-19 outcomes between counties with IHEs that *reported any campus testing* and counties with IHEs that *reported no campus testing*.

**Figure 1:**
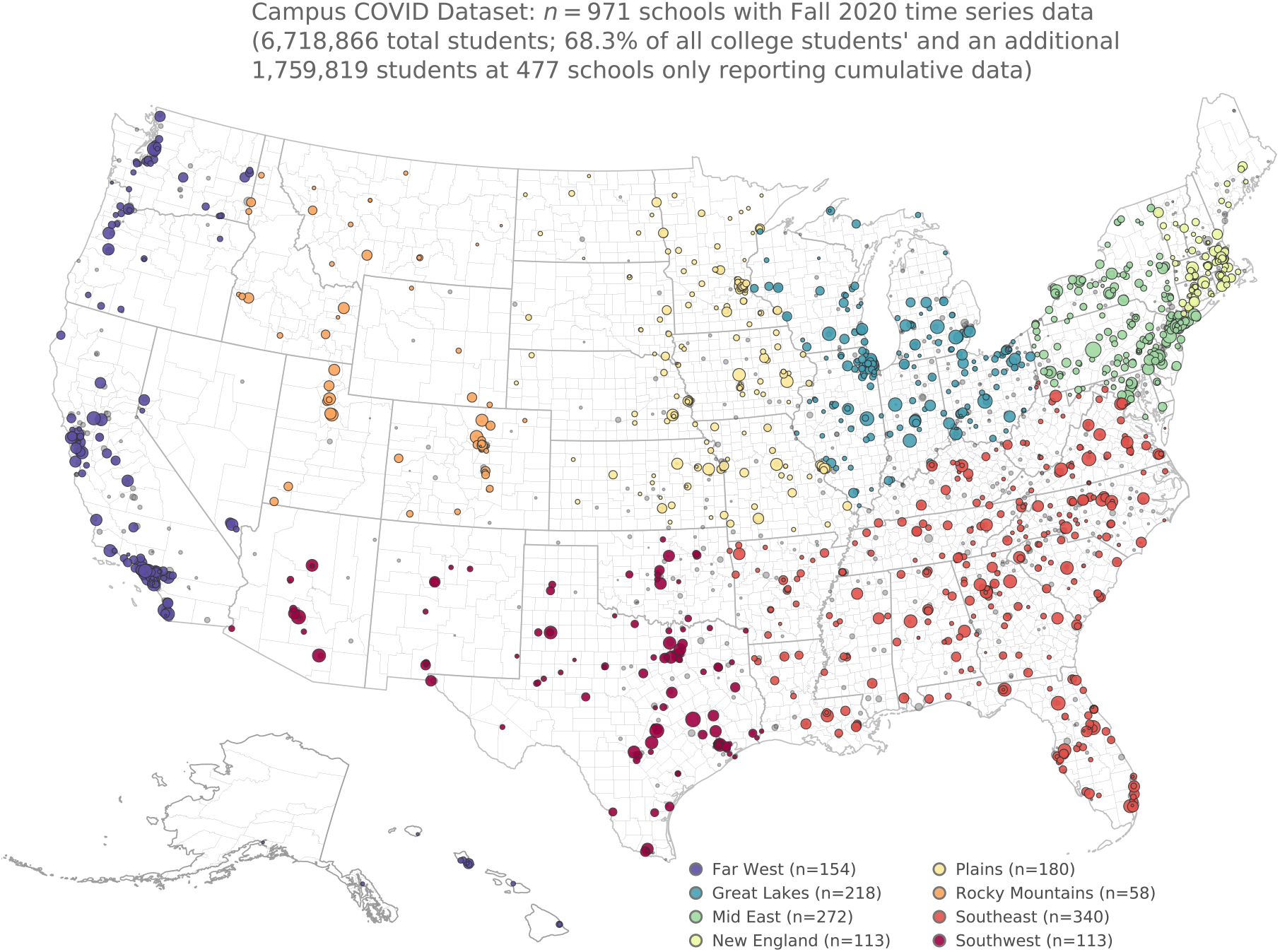
Description of the Campus COVID Dataset. Map of the 1,448 institutes of higher education included in the Campus COVID Dataset. The dataset includes semester-long time series for 971 institutes of higher education (see SI A.4 for several examples), in addition to 477 that have cumulative data only (i.e. one sum for the total testing and/or case counts for the Fall 2020 semester).

COVID-19 has had a disproportionate impact on older populations, and we see especially high death rates in regions with more congregate senior living and long-term care facilities [34, 35]. On the other hand, regions with more young people (i.e., “college towns”—or, here, college counties) experienced relatively fewer hospitalizations and deaths [35]. This means that care should be taken when comparing averages between groups of counties, and prior to creating the groups, we must attempt to match the underlying socio-demographics of the study groups. In the following, from our dataset with more than 1,238 different counties with IHEs, we generate two matched groups of counties—those with IHE students who returned to the Fall 2020 semester primarily in-person (*n*_*A*_ = 393 total) and those with IHE students who remained primarily online (*n*_*B*_ = 449 total) with as similar distributions of demographics as possible. The matching procedure, described in details in the Supplementary Information (SI), considers on five county socio-demographic variables: age, race, income, total population, and urban-rural code. The resulting groups were spatially heterogeneous, and did not ultimately include counties from regions that experienced early surges in March, 2020 (e.g., counties in New York City, etc; see SI A.2), which could have confounding effects.

## Fall 2020 reopening status: In-person vs. online education

With the two matched groups, we compare the average new cases and new deaths per 100,000 between the two groups. In Figure 2a, we see that during July and August the number of new cases per 100,000 was almost identical for the in-person and online counties. By the end of August (i.e., the start of the Fall 2020 semester), we begin to see these two curves diverge; college counties with primarily in-person enrollment report more new cases per 100,000 on average for the remainder of 2020, a gap that narrows shortly after the Fall 2020 semester ends (cumulative cases per 100,000 during the study period (August to December, 2020) among: counties with primarily online IHEs = 4,196.3, 95% CI: [4,042.9 – 4,347.6]; counties with primarily in-person IHEs = 5,415.3, 95% CI: [5,310.6 – 5,552.0]). We see the same trend—but lagged by about four weeks—when comparing the average new deaths per 100,000 between the two groups of counties. These analyses, even after controlling for several potentially confounding socio-demographic variables, highlight clear differences in COVID-19 outcomes based on IHE reopening policy (cumulative deaths per 100,000: counties with primarily online IHEs = 44.5, 95% CI: [45.4 – 48.1]; counties with primarily in-person IHEs = 67.2, 95% CI: [64.1 – 72.0]). In SI A.2, we also show how the matching procedure used here excludes other potentially confounding *spatial* variables as well. For example, counties that were hit early and hard by COVID-19 in March and April of 2020 (e.g. counties in New York City, greater Boston area, etc.) are already not included in these averages (see a visualization of the included counties in Figure A.3).

**Figure 2:**
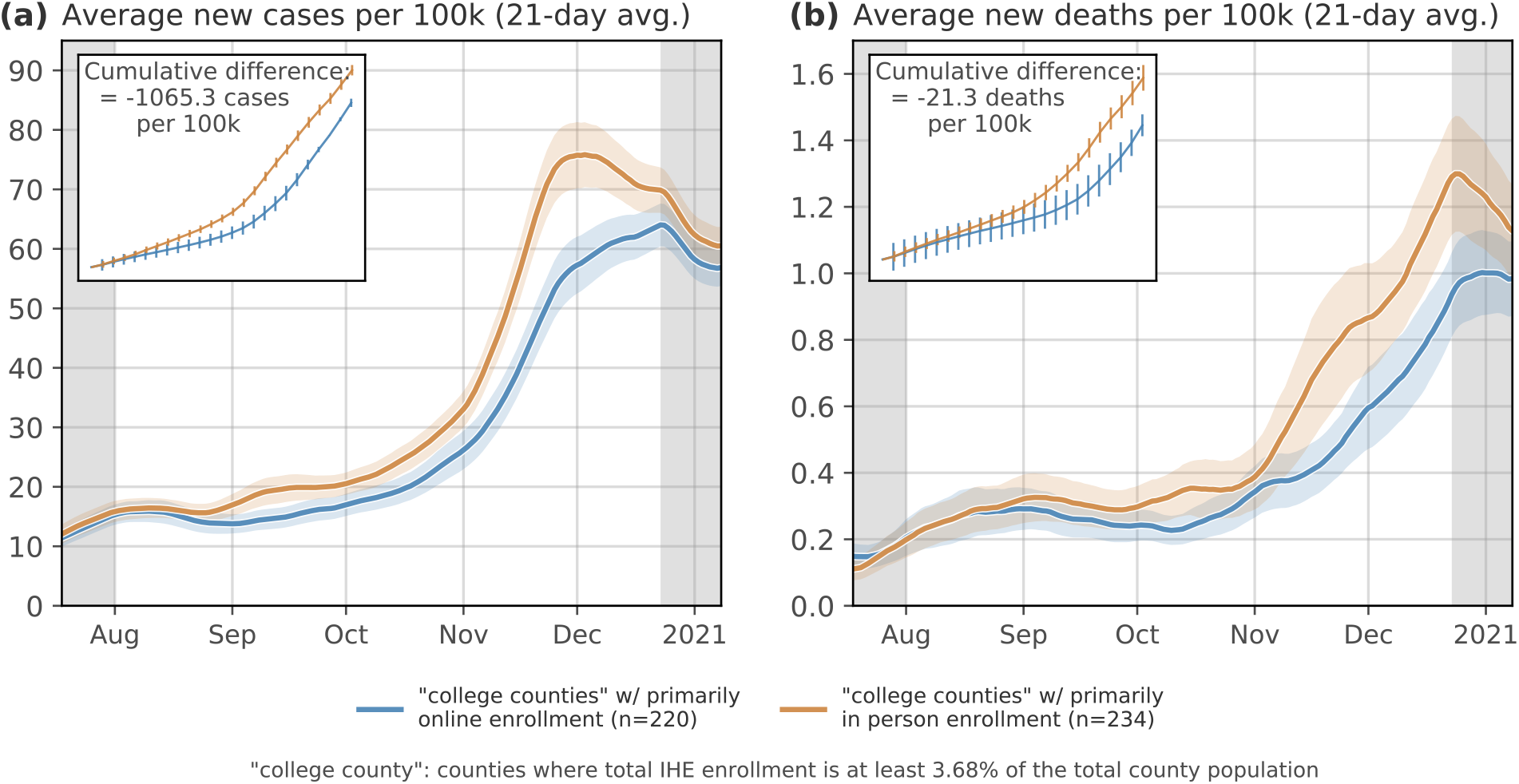
Counties with IHEs categorized as in-person vs. online for Fall 2020. Here, we compare the average **(a)** new cases and **(b)** new deaths per 100,000 in counties with IHEs that were categorized as “primarily in-person” vs. “primarily online” for the Fall 2020 semester. IHEs classified with “hybrid” reopening strategy were not included in this comparison as there is a great deal of heterogeneity in what constitutes a “hybrid” reopening. IHE reopening data is from [23]. Ribbons: 95% confidence interval. Insets: Cumulative differences between counties primarily in-person/online IHEs. Cumulative cases per 100,000 among primarily online counties during the study period (August - December, 2020) = 4,196.3, 95% CI: [4,042.9 – 4,347.6]; primarily in-person counties = 5,415.3, 95% CI: [5,310.6 – 5,552.0]. Cumulative deaths per 100,000 among primarily online counties = 44.5, 95% CI: [45.4 – 48.1]; primarily in-person counties = 67.2, 95% CI: [64.1 – 72.0].

## Quantifying the benefits of IHE-affiliated testing

The extent to which IHEs tested their students and employees for COVID-19 varied substantially: some schools focused their limited testing resources on only testing symptomatic individuals while others developed a strict and massive testing program that required frequent (e.g. weekly) asymptomatic testing. Because of this heterogeneity, we sought the simplest distinction for comparing groups of counties; we split the “primarily in person” counties into two groups of counties: those with IHEs that reported conducting *any* COVID-19 tests on campus and those that reported *none*. Of the *n* = 234 “primarily in person” counties, the Campus COVID Dataset includes data from *n* = 144 counties. Often, when IHEs do not administer any on-campus tests, they have a form for students and staff to self-report results from external testing providers (e.g. pharmacies, health clinics, etc.). In order to classify an IHE as “non-testing” we sought out official documentation on the IHE’s websites, though a key limitation of this approach is that an IHE could have been conducting testing without posting updates to their websites.

Counties with IHEs that reported conducting a nonzero number of tests saw, on average, a similar amount of cumulative cases during the Fall 2020 semester but significantly fewer deaths (Figure 3; inset subplots show weekly average cumulative cases and deaths). Notably, we see an increase in the number of cases per 100,000 on average in early September 2020 among counties with IHEs that do report testing (i.e., the campus testing is working as designed—detecting cases in the campus population; Figure 3a); this same increase in reported cases *does not* appear among counties with IHEs that do not report testing. This suggests that the return-to-campus surges that were being detected in IHEs that report testing may have occurred but remained undetected or under-reported in counties without IHE-affiliated testing. This suggestion is in part corroborated by the increase in deaths in the middle of October among counties without IHE testing, which does not appear to follow a commensurate increase in case counts (Figure 3b). Additionally, because the two groups of counties are well-matched along demographic variables that influence COVID-19 related deaths, we would not expect relatively similar case rates between the groups to yield significantly different death rates, though this is what we see (cumulative deaths per 100,000 among: counties with IHE testing = 56.0, 95% CI: [54.6 – 60.9]; counties with no reported IHE testing = 70.2, 95% CI: [64.4 – 81.0]).

**Figure 3:**
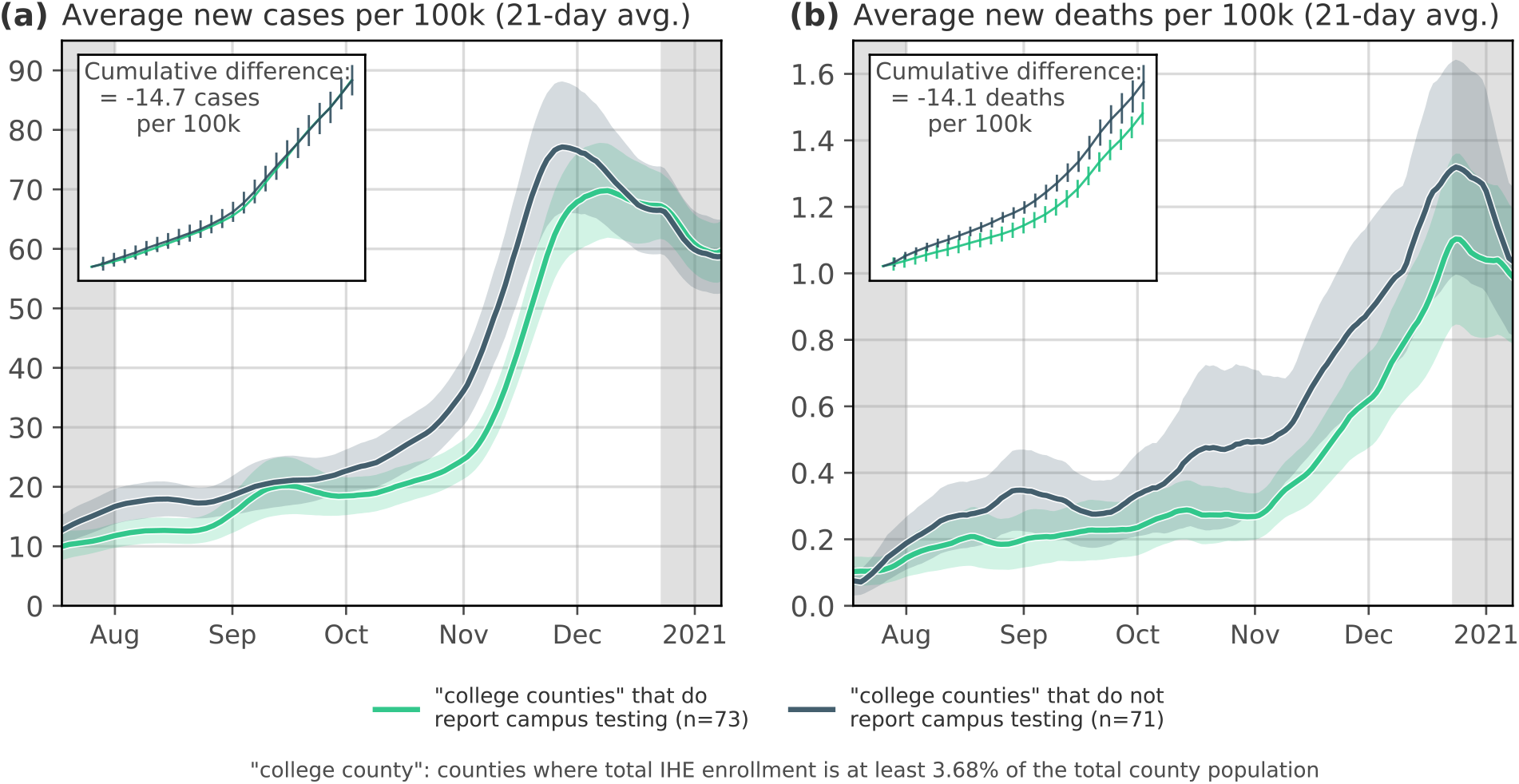
Comparing counties with IHEs that reported any vs. zero COVID-19 tests. As in Figure 2, we compare the average **(a)** new cases and **(b)** new deaths per 100,000 in counties with IHEs reported conducting any COVID-19 tests vs. counties with IHEs that reported no tests. Note: if there are multiple IHEs in a single county, we sum together the total number of tests between all IHEs. Ribbons: 95% confidence interval (CI). Insets: Cumulative differences between counties with/without testing IHEs. Cumulative cases per 100,000 among testing counties during the study period = 5,235.7, 95% CI: [5,138.5 – 5,315.6]; non-testing counties = 5,264.1, 95% CI: [4961.0 – 6,009.7]. Cumulative deaths per 100,000 among testing counties = 56.0, 95% CI: [54.6 – 60.9]; non-testing counties = 70.2, 95% CI: [64.4 – 81.0].

Importantly, while the two groups of counties used in this comparison were relatively balanced with respect to socio-demographic variables, they are not formed based on information about differences in county-level mitigation policies that may have been active in the counties during this time period. For this reason, we conducted additional analyses where we used state-level policy data [36] to quantify the effect of IHE testing policy while controlling for a number of county-level socio-demographic variables as well as the number of active mitigation policies in place. In SI A.3 we report the results of Generalized Linear Model regression to quantify the extent to which this time series of policy measures—along with data about IHE testing and enrollment policy, demographic data about the county itself, and average temperature—explains COVID-19-related deaths (Table A.4).

The results of this model again highlight the importance of IHE testing, while also controlling for other common confounding variables. In Table A.4, we see that the “IHE testing” variable has a significant negative correlation to the (lagged) number of deaths per 100,000, whereas the tests run at the county level are positively correlated with deaths. This is understandable since the amount of asymptomatic testing taking place during the Fall 2020 semester of individuals *not* affiliated with IHEs is quite low [37]. The model also reports a negative correlation with average temperature, which is consistent with previous research [38] and is broadly intuitive: as winter approaches, people in colder counties may be more likely to spend their time indoors, which may facilitate easier transmission of the virus. The model also finds the expected importance of control variables such as the median income variable (negative correlation with deaths) and population over 60 (positive correlation with deaths).

## Case study: Higher education in Massachusetts

According to the data collected in this work, IHEs in Massachusetts administered more COVID-19 tests to students and staff, on average, than most other states. As such, the Campus COVID Dataset includes time series data for 56 IHEs during both the Fall 2020 and Spring 2021 semesters. Additionally, the Massachusetts Department of Health releases weekly data about testing and case counts at the *city* level (total of *n* = 351 cities instead of *n* = 15 counties) [39]. In this section, we analyze Massachusetts as an informative case study about the role that IHE-affiliated testing may play in a community’s response to COVID-19.

In February, 2021—at the start of the Spring semester—the University of Massachusetts, Amherst (UMass Amherst) experienced a large COVID-19 outbreak among the campus community. Throughout the Fall 2020 semester, UMass Amherst followed a campus testing regimen that required frequent testing of on-campus students and staff; as a result, the city of Amherst’s average number of tests per 1,000 residents was far higher than that of other cities in Massachusetts (Figure 4c). After the Fall 2020 semester ended, the overall testing volume in Amherst sharply declined since the number of students on campus decreases during December and January; the timing of this decline coincided with large *increases* in COVID-19 cases both regionally and statewide (Figure 4b & 4c). However, as neighboring cities to Amherst began to report a surge in cases during this period (Figure 4b), the city of Amherst did not report a commensurate rise in cases. It is possible that there were simply not as many cases in Amherst during December and January, but because there was such a large decrease in the amount of tests conducted during that period, it is also possible that there were some infections that remained undetected.

**Figure 4:**
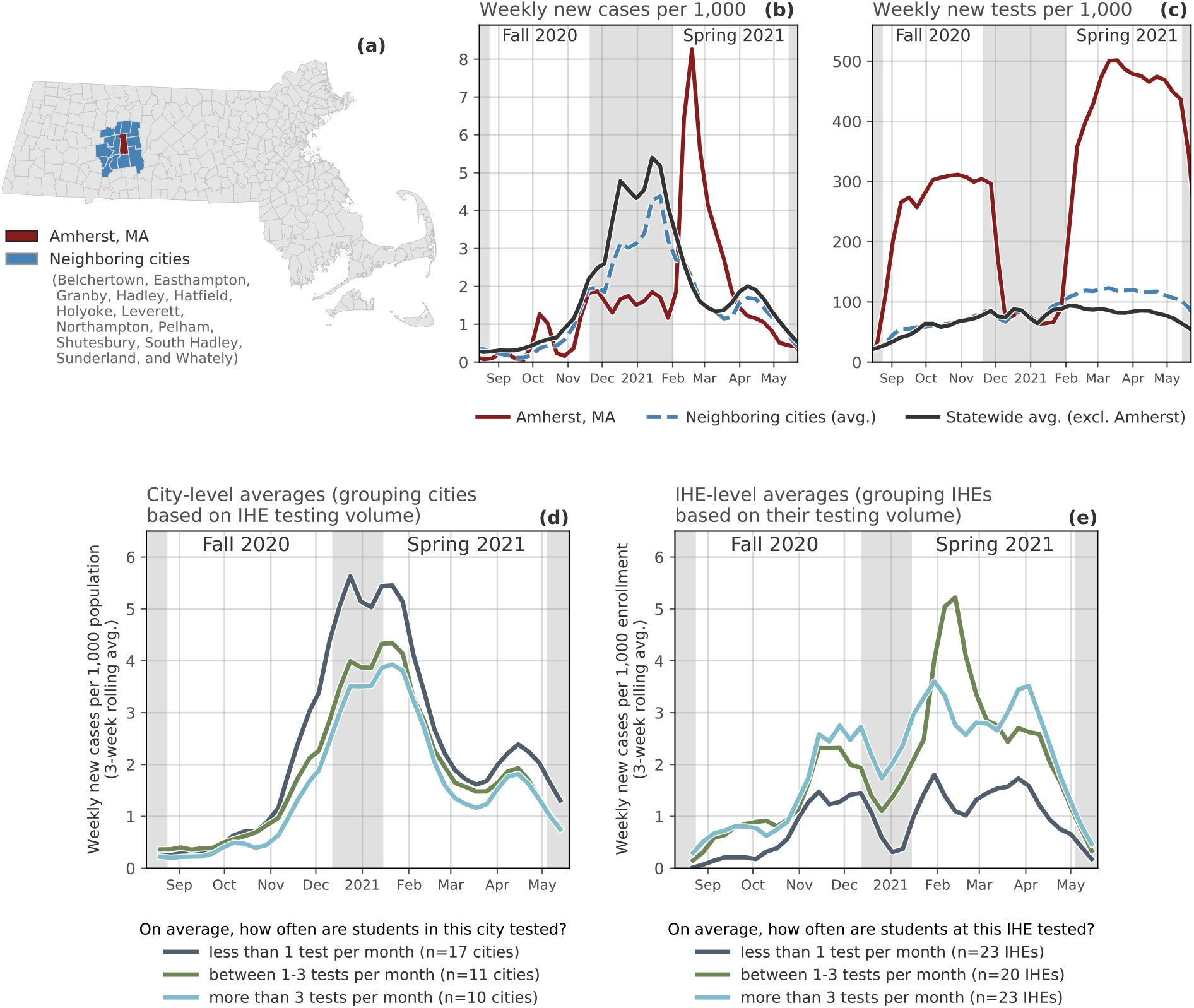
Case Study: COVID-19 in Massachusetts college cities. Top: High-lighting testing and case counts in and around Amherst, Massachusetts. **(a)** Map of Massachusetts cities; in this map, the city of Amherst is red and the surrounding cities are colored blue. **(b)** Time series of weekly new cases per 1,000 in: Amherst, the surrounding cities, and the rest of Massachusetts. **(c)** Time series of weekly new tests per 1,000 in: Amherst, the surrounding cities, and the rest of Massachusetts. Bottom: Comparing outcomes of cities and IHEs with more/less IHE-affiliated testing. **(d)** City-level average weekly new cases per 1,000, grouped by cities with IHEs that test students on average fewer than once a month, between one and three times a month, and over three times a month (Note: we sought out city-level data for COVID-19 deaths, but the state does not report these). **(e)** IHE-level average weekly new cases, grouped by IHEs that test students on average fewer than once a month, between one and three times a month, and over three times a month.

Either way, when students returned to campus in January, they returned to a city with a testing rate that was lower than it had been in late November, 2020. During the first few weeks of the Spring 2021 semester, UMass Amherst reported almost 1,000 new infections among students and staff, one of the largest outbreaks in the country at that time [40]. It is difficult to know whether the UMass Amherst outbreaks were primarily the result of importation from different areas following students’ return to campus or whether the returning students instead became infected following interactions with Amherst residents (or both). Regardless the source of these cases in Amherst, what happened *after* the February surge highlights the role that IHEs can have in local mitigation; testing volume in Amherst increased dramatically during the Spring 2021 semester, students who tested positive were strictly isolated, and on-campus restrictions of activities were put in place [41]. The example of UMass Amherst is a useful case study for highlighting a broader trend among Massachusetts cities with IHEs—a trend that resembles the results in from our analyses of testing. In Figure 4d we show the average new cases per 1,000 for cities with IHEs that test their students an average of a) less than once per month, b) between 1-3 times per month, and c) more than three times per month; on average, cities with IHEs that conduct *more* tests also have *fewer* new cases. This pattern does not hold when only looking at on-campus cases from IHEs (as opposed to citywide cases); instead, we see that IHEs that test less also report fewer cases (Figure 4e). This trend may emerge because low-testing IHEs are not conducting enough tests to detect the true number of cases on campus, though proving this definitively is almost impossible without detailed contact tracing and/or retrospective antibody testing.

## Discussion

The COVID-19 pandemic required governments and organizations to implement a variety of non-pharmaceutical interventions (NPIs) often without a thorough understanding of their effectiveness. Policy makers had to make difficult decisions about which policies to prioritize. While a body of literature has emerged since the beginning of the pandemic about measuring the effectiveness of NPIs [42–45], to date there have been no studies that attempt to measure the effectiveness of campus testing systematically nation wide. This study sheds light on this topic by directly measuring the impact of campus testing on county level COVID-19 outcomes. We collected data from 1,448 colleges and universities across the United States, recording the number of tests and cases reported during the Fall 2020 semester; by combining this data with standardized information about each school’s reopening plan, we compared differences in counties’ COVID-19 cases and deaths, while controlling for a number of demographic variables.

When looking at county COVID-19 outcomes, our results shows that COVID-19 outcomes were worse in counties with IHEs that report no testing and in counties where IHEs returned to primarily in-person instruction during the Fall 2020 semester. These findings support the CDC recommendation to implement universal entry screening before the beginning of each semester and serial screening testing when capacity is sufficient [46] and are in line with smaller scale, preliminary results from other studies [47, 48]. It is worth stressing that the matched groups selected here (no reported testing vs. any reported testing) do not provide insights into the ideal amount of IHE testing needed to manage campus outbreaks. However, we examine this question briefly by grouping cities in Massachusetts based on the amount of IHE testing, as opposed to simply whether they have IHE testing or not. Future work will examine whether there are optimal trade-offs between testing volume, cost of testing, levels of local transmission, and community demographics [49]. Furthermore, this study does not look at optimal testing strategies, it offers evidence for the protective effect of campus testing in any form and reopening status on county COVID-19 outcomes.

The COVID-19 pandemic highlighted the importance of data standardization for understanding the impact of the virus but also in to inform response, resource allocation, and policy. While much attention has been given to this topic for data reported by healthcare and public health organizations, little attention has been given for COVID-19 case and testing data reported by IHEs. A significant portion of the effort undertaken by this study was spent compiling and standardizing the data across IHEs nationwide. In the cases where IHEs did report campus testing data, the ease of access varied widely and oftentimes different metrics for cases and testing were reported out. For example, some IHEs would report only active cases, cumulative cases, or number of isolated individuals. Similarly, sometimes there would be no distinction between types of test given or temporal information on when the test was given. In their campus testing guidance [46], the CDC should also include recommendations on data standards and reporting formats.

Although the availability of vaccine is a game changer in our approaches at controlling SARS-CoV-2 infections, concerns remain around lingering outbreaks caused by new variants emerging, ongoing transmission in the rest of the world, vaccine hesitancy, and the possibility of waning effectiveness of the current vaccines [50, 51]. In regions like the Mountain West and South, vaccination rates remain disproportionately low among younger adults and the general population when compared to nation wide averages [52]. States in these same regions are also disproportionately represented among the states with the lowest IHE testing in our dataset. Given the number of younger adults enrolled in IHEs, the increased mobility and international nature of this population, and the fact that this population is less likely to practice COVID-19 mitigation behaviors, our study suggests that campus testing in counties with a substantial student population can play a relevant role in mitigating the number of COVID-19 outbreaks in the surrounding population.

## 2 Data & Methods

### 2.1 Data collection and sources

County-level case data are from the COVID-19 Data Repository by the Center for Systems Science and Engineering (CSSE) at Johns Hopkins University [37]. County-level population and demographic data are from the 2018 American Community Survey (ACS) [53]. Weekly data for testing and case counts in Massachusetts cities are from the Massachusetts Department of Public Health [39]. Data about IHEs—including the number of full-time students and staff, campus location, institution type, etc.—come from the Integrated Postsecondary Education Data System (IPEDS) via the National Center for Education Statistics [54].

Data about individual IHEs’ plans for returning to campus (i.e., online only, in-person, hybrid, etc.) come from the College Crisis Initiative at Davidson College [23]. This dataset classifies IHEs based on the following categories, which we use to create three broader categories (in parentheses): “Fully in person” (primarily in-person), “Fully online, at least some students allowed on campus” (primarily online), “Fully online, no students on campus” (primarily online), “Hybrid or Hyflex teaching” (hybrid), “Primarily online, some courses in person” (primarily online), “Primarily in person, some courses online” (primarily in person), “Primarily online, with delayed transition to in-person instruction” (primarily online), “Professor’s choice” (hybrid), “Simultaneous teaching” (hybrid), “Some of a variety of methods, non-specific plan” (hybrid). We did not include “hybrid” IHEs in our analyses here, but they remain an interesting avenue for future research, which we strongly encourage using the Campus COVID Dataset.

### 2.2 The Campus COVID Dataset

The Campus COVID Dataset was collected through a combination of web scraping, manual data entry, or communication with administrators at IHEs. In sum, the process involved collecting thousands of URLs of the COVID-19 dashboards (or analogous website) of each of over 4,000 IHEs, which we then used for manual data collection, inputting time series of case counts and testing volume between August 1 and December 16, 2020. The data for each IHE is stored in its own Google Sheet (indexed by a unique identifier, its ipeds_id), the URL of which is accessible through a separate Reference sheet. For full details on the data collection process, see SI A.1.

## Data Availability

The dataset and Python code to reproduce the analyses and construction of the database is available at https://github.com/jkbren/campus-covid.

## Additional information

### Software and data availability

The dataset and Python code to reproduce the analyses and construction of the database is available at https://github.com/jkbren/campus-covid [33].

## Acknowledgements

The authors thank Kaitlin O’Leary, Representative Mindy Domb, Timothy LaRock, Daniel Larremore, Maciej Kos, Jane Adams, Rylie Martin, Addie McDonough, Anne Ridenhour, Benjy Renton, and Mike Reed for helpful discussions and additions to the dataset. A.V. and M.C. acknowledge support from COVID Supplement CDC-HHS-6U01IP001137-01 and Cooperative Agreement no. NU38OT000297 from the Council of State and Territorial Epidemiologists (CSTE). A.V. acknowledges support from the Chleck Foundation. N.G. acknowledges LA-UR-21-25928. The findings and conclusions in this study are those of the authors and do not necessarily represent the official position of the funding agencies, The Rockefeller Foundation, the National Institutes of Health, or U.S. Department of Health and Human Services.

## Author contributions

B.K. and N.G. created the data collection protocol for the Campus COVID Dataset and collected the data with Z.B, B.L, R.G, P.K.; some of these data were collected through a crowd-sourced survey. C.M. directed the College Crisis Initiative, which is the source of campus reopening data. B.K. and N.G. performed the analyses. B.K., S.M., M.C., J.G., D.L., C.R.M., S.V.S., and A.V. guided the research. All authors contributed to and/or approved the manuscript.

## Citation diversity statement

To evaluate gender bias in the references used here, we obtained the gender of the first/last authors of the papers cited here through either 1) the gender pronouns used to refer to them in articles or biographies or 2) if none were available, we used a database of common name-gender combinations across a variety of languages and ethnicities. By this measure (excluding citations to datasets/organizations, citations included in this section, and self-citations to the first/last authors of this manuscript), our references contain 12% woman(first)-woman(last), 21% woman-man, 22% man-woman, 38% man-man, 0% nonbinary, 4% man solo-author, 3% woman solo-author. This method is limited in that an author’s pronouns may not be consistent across time or environment, and no database of common name-gender pairings is complete or fully accurate.

