## Supplementary Information for "Higher education responses to COVID-19 in the United States: Evidence for the impacts of university policy"

### A Supplemental Information

#### A.1 The Campus COVID Dataset

##### A.1.1 Collecting URLs of COVID-19 dashboards

We used the Python library `googlesearch` [1] to automate Google search queries, and we used the `gspread` Python library [2] to aggregate the output into a single Reference Google Sheet. For each of over 4,000 IHEs in the IPEDS 2018 data, we generated the following Google search queries: "`university_name` COVID dashboard", "`university_name` COVID cases", "`university_name` COVID testing", with and without quotation marks to increase specificity or broaden the search yields (if the IHE's name does not include the state it is located in, we append the state name to the search queries to avoid duplicate or similarly-named institutions in different states). This process produced several candidate URLs, and we select the most common URL to include in the Reference Google Sheet; this was usually effective at creating one single URL for each IHE, since "COVID cases" and "COVID testing" are semantically similar terms that are likely to co-occur on the COVID-19 information pages for universities. We repeated this process for thousands of IHEs, and we were left with URLs for the COVID-19 dashboards (or analogous website) of 2,739 IHEs.

##### A.1.2 Standardized data collection

In order to create a more streamlined data collection process, we created separate pre-formatted Google Sheets for every IHE using the `gspread` Python library. Each of these Google Sheets was indexed by the `ipeds_id` of the IHE and was formatted like Table A.2. The "total_tests" and "positive_tests" columns were left blank and eventually filled in by research assistants, who would visit the COVID-19 dashboard urls of each IHE and record the data. Many IHEs elected to report data using interactive visualization software, such as Microsoft Power BI or Tableau dashboards; others used a simple data table to report their numbers; others wrote weekly email updates to the campus community that included the testing and case counts for the previous week.

Many IHEs reported none of this information, for a variety of potential reasons. For one, many IHEs were closed for the Fall 2020 semester (or entirely, due to financial hardship), not allowing students on campus at all and therefore not actively recording case counts among the campus population. Some schools were at least partially in-person for the Fall 2020 semester but did not appear to collect data about cases on campus. We categorized these schools as "cannot find data". Other IHEs had passages of text on their websites that suggest they had knowledge of the number of cases on campus (e.g. "case counts on campus are low") but did not publicly report raw numbers. These schools were categorized as "not publishing data". These categorizations can be found in Table A.1.

| | |
|---|---|
| complete time series | 971 |
| cumulative data only | 477 |
| cannot find data | 1,271 |
| total IHEs in data | 1,448 |
| total IHEs searched | 2,719 |

**Table A.1: Current status of the Campus COVID Dataset.** In total, the Campus COVID Dataset includes data about more than 1,400 IHEs. To collect these data, we searched among over 2,719 IHEs; approximately 40% of these are IHEs with data that we could not find (because the IHE does not collect self-reported positive tests and/or does not conduct campus testing, etc.) or with data that we believe exists but was not being shared publicly by the IHE. There are over 971 IHEs with time series of testing and/or case counts for the Fall 2020 semester. If an IHE reported only cumulative testing or case counts, we classify it as "cumulative only".

| date | total_tests | positive_tests | college | URL | ipeds_id | notes |
|---|---|---|---|---|---|---|
| 2020-08-01 | | | `university_name` | `dashboard_url` | `ipeds_id` | |
| 2020-08-02 | | | `university_name` | `dashboard_url` | `ipeds_id` | |
| 2020-08-03 | | | `university_name` | `dashboard_url` | `ipeds_id` | |
| ... | ... | ... | ... | ... | ... | |
| 2020-12-16 | | | `university_name` | `dashboard_url` | `ipeds_id` | |

**Table A.2: Example template for inputting data.** Each IHEs in the Campus COVID Dataset has a unique URL that leads to a dataframe with this structure. For each date that the IHE reports a number of new cases ("positive_tests" above) or new tests administered ("total_tests" above), we input that value in its corresponding row. For IHEs that report testing and case counts weekly, we insert the data at the first collection date, which makes for more accurate smoothing when performing 7-day averages. If the IHE only reports *cumulative* cases or tests for the Fall 2020 semester, we leave the "total_tests" and "positive_tests" columns blank and report the "cumulative_tests" and "cumulative_cases" in the "notes" column, which we extract later in the analyses.

#### A.2 Matching counties with broadly similar demographics

Because COVID-19 outcomes correlate with demographic variables (e.g. age [3], among others), extra care must be taken when comparing averages between groups of counties to ensure that any observed differences are not due to differences in population structure of the underlying populations. Here, we try to create counties with as similar as possible distributions of age, race, income, urban-rural code, and population size. (Note: "urban-rural code" is an ordinal variable ranging from 1 ("large central metro") to 6 ("non-core") and is assigned by the National Center for Health Statistics [4].)

In order to compare differences in average case counts and deaths (as in Figure **??**) among counties with IHEs that returned to in-person education compared to those with IHEs that remained online, we rely on data collected by the College Crisis Initiative [5].

This dataset assigns a category for over 1,800 IHEs based on the reopening strategy for the Fall 2020 semester. From these IHEs, we sum together the total full-time enrollment in each reopening category (e.g. a county may have three universities—with 250, 800, and 15,000 students enrolled full-time; if the school with 15,000 students is categorized as "primarily online" and the other two are "primarily in-person" we say that specific county has 15,000 primarily online students and 1,050 primarily in-person students). As discussed in the main text, the final piece for constructing groups of counties with similar demographics is the percent of the county population affiliated with the IHE. That is, we define a "college county" based on the what percent of the total population is made up of students enrolled full-time at an IHE in the county.

We create two groups of counties from the 1,238 unique counties in the College Crisis Initiative [5]; we iteratively vary the threshold of inclusion into these two groups, finding the threshold that minimizes the total Jensen-Shannon Divergence (JSD; the JSD between two distributions, $P$ and $Q$, is $JSD(P||Q) = \frac{1}{2}D(P||M) + \frac{1}{2}D(Q||M)$, where $M = \frac{1}{2}(P + Q)$ and $D(P||Q) = \sum_{x \in X} P(x) \log(\frac{P(x)}{Q(x)})$, the Kullback-Leibler Divergence) between the average demographic distributions of the counties that comprise the groups. In our case, this threshold corresponds to the percent of the total county population enrolled in IHEs within that county full-time. Intuitively, a "college county" is one with a relatively large percentage of its population being affiliated to an IHE, but the precise value of this percentage is not commonly defined. Here, we test a range of thresholds and select the one that minimizes the JSD between the resulting groups' distributions of the five demographic variables of interest (see Figure A.1).

By selecting the threshold that minimizes the average JSD between the demographic distributions of the two groups, we get closer to making sound comparisons along the true axis of interest. Following this procedure, the resulting groups of counties are similar in demographics (Figure A.2), spatially spread out across the country, and minimally clustered based on region (Figure A.3).

#### A.3 Statistical controls for counties with active mitigation policies

While the two groups of counties—the "primarily in-person" vs. "primarily online" counties—are broadly similar across demographic categories (Figure A.2), there could still be underlying differences between the two groups that influence their different COVID-19 outcomes. For example, this could happen if the two groups differed in the extent to which they enacted mitigation policies (i.e., if there were a common variable influencing whether a given county introduced mitigation policies as well as whether IHEs in the county remained primarily online vs. in-person during the Fall 2020 semester). There are a number of possible sources of this variability, ranging from differences in population density [6], to differences in messaging from political leaders [7]. In the model below, we include the data about voting patterns in the 2020 presidential election in order to control for potential biases arising from differences in political behavior at the county level.

To control for potential biases arising from differences in local mitigation policies, we

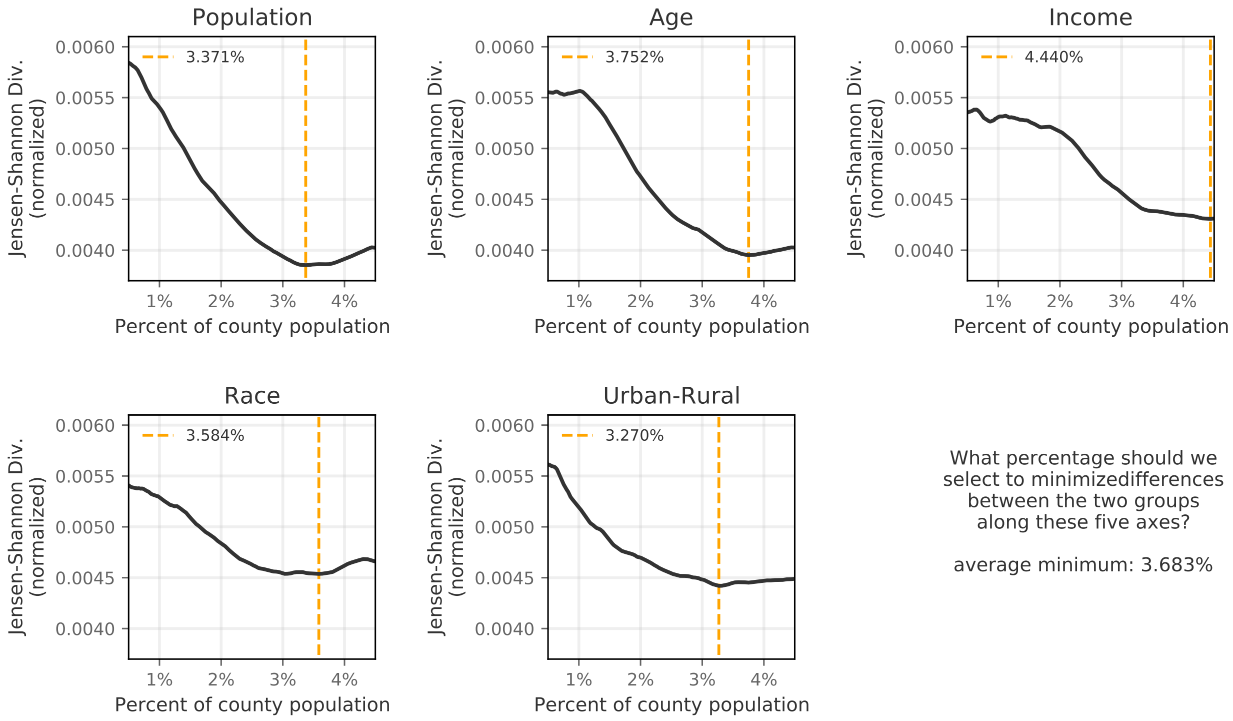

**Figure A.1: Jensen-Shannon Divergence between distributions of demographic variables.** As we vary the threshold for inclusion into the two groups—counties with IHEs that returned primarily in-person for Fall 2020 and counties with IHEs that remained primarily online—the Jensen-Shannon Divergence also changes. We want to select the value for this threshold based on whatever minimizes the Jensen-Shannon divergence, on average.

assigned each county to an "active mitigation policies" score based on policy tracking data from the Oxford COVID-19 Government Response Tracker [8]. These are daily time series data indicating whether or not a number of different policies were active on each day for a given state. Not only does this dataset list the presence or absence of a given policy, it also includes information about the severity (e.g. restrictions on gatherings of 10 people vs. restrictions on gatherings of 100 people, or closing all non-essential workplaces vs. closing specific industries, etc.). From these indicator variables, Hale et al. (2021) define a summary "stringency index" that characterizes the daily intensity of the mitigation policies that a given region is undergoing over time. We include this "stringency index" variable in an Generalized Linear Model regression to quantify the extent to which this time series of policy measures—along with data about IHE testing and enrollment policy, demographic data about the county itself, and average temperature—predicts COVID-19-related deaths (Table A.4). The descriptions of the variables used in the regression can be found in Table A.3 and are visualized in Figure A.4.

This model predicts deaths per 100,000 county population at a 38-day lead time (i.e., using data from today to predict the number of COVID-19 deaths reported 38 days from

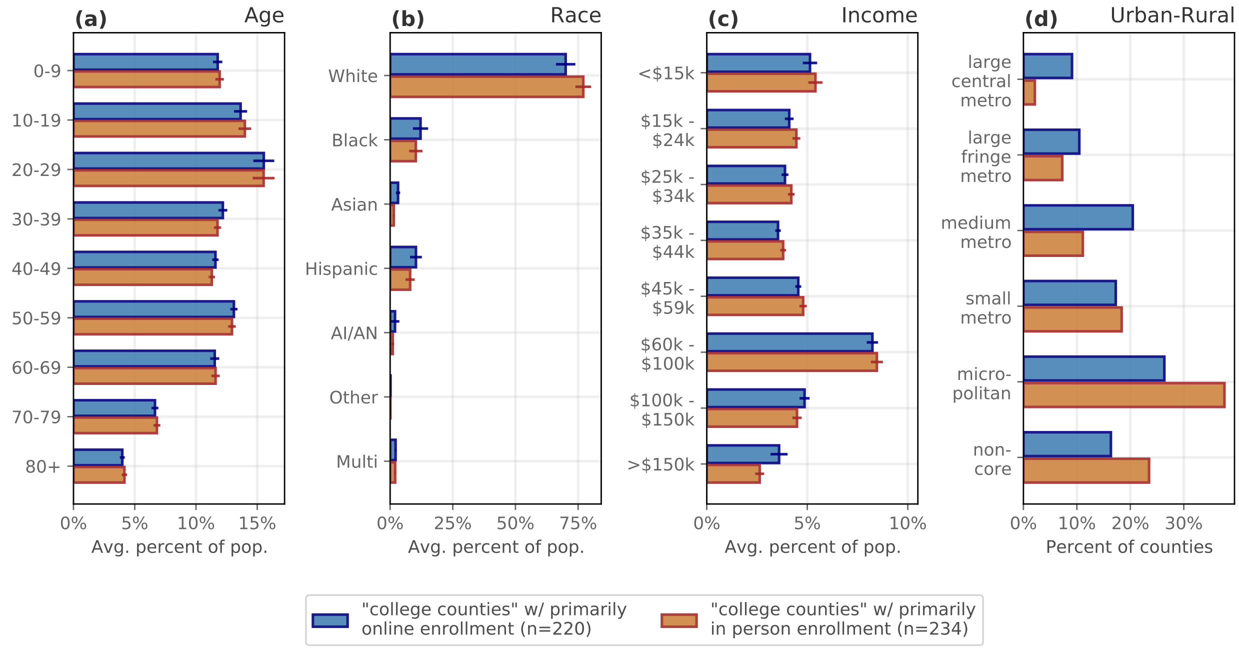

**Figure A.2: Comparison of county-level demographics between groups.** Here, we compare the two groups—counties with IHEs that returned primarily in-person for Fall 2020 and counties with IHEs that remained primarily online—based on distributions of **(a)** age, **(b)** race, **(c)** income, and **(d)** urban-rural designation. Error bars: 95% confidence intervals.

now), which was selected because it is the lag that maximizes the Pearson $\chi^2$ of the model. Note: this specific value, 38 days, is in line with the CDC's median window of the time between the onset of infection and death [14], and by varying the lead time we do not see any substantial differences in the sign, value, and significance of the coefficients for the different variables. To test for possible multicollinearity, we assigned a Variance Inflation Factor ($VIF$) to each of the variables in regression. The $VIF$ is defined as $VIF = 1/(1-R_i^2)$; a $VIF = 1$ indicates that a variable is uncorrelated, and $VIF$ between 5-10 suggests that a given variable is highly correlated in the regression. For the regression from Table A.4, the $VIF$ values are: "average temperature" = 1.226, "urban/rural code" = 1.951, "population density" = 2.244, "median income" = 1.422, "population over 60" = 1.408, "2020 voting behavior" = 1.753, "IHE fulltime enrollment" = 1.492, "IHE fulltime enrollment (online)" = 4.554, "IHE fulltime enrollment (in person)" = 4.042, "stringency index" = 1.094, "county new tests" = 1.408, "IHE new tests" = 1.498.

There are limitations to the regression used here. For one, the policy data we use is at the state level, while the analyses are at the county level. Part of the reason for this is that there remains inconsistent data about county-level policies and more reliable data at the state level. Also, among datasets that do include finer-scale policy data (e.g. [15]), it is often about specific cities, as opposed to counties. Another limitation of using these kinds of policy data in our analysis is that we still do not have information about adherence to

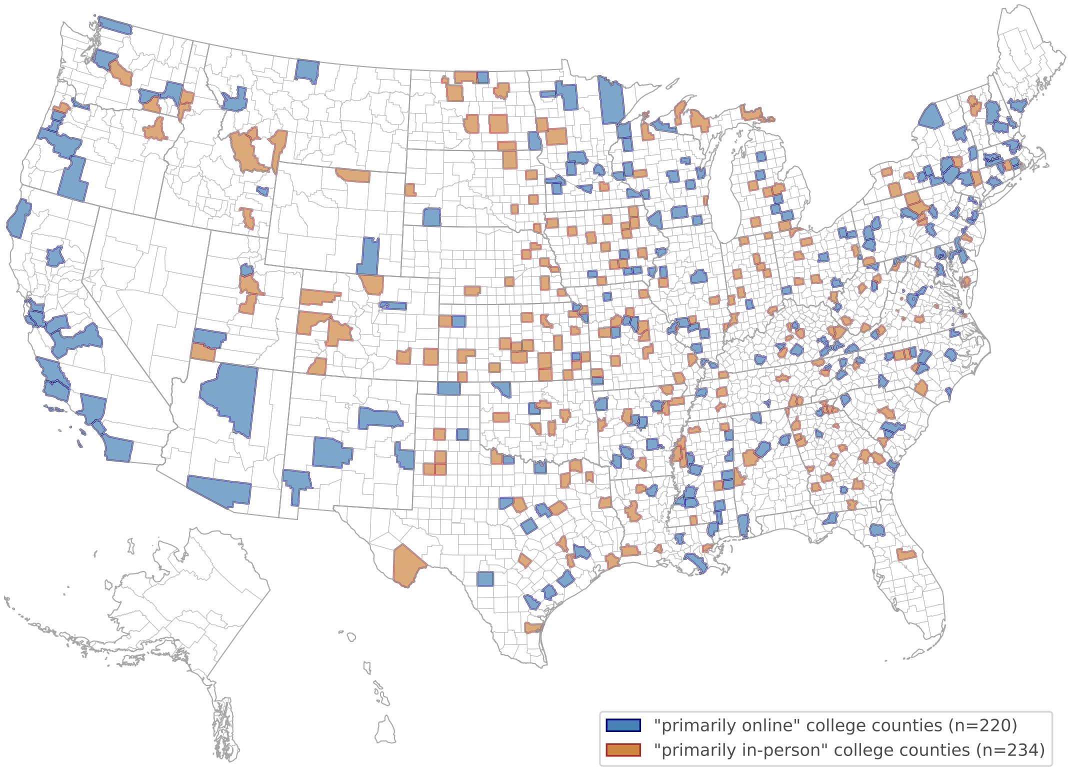

**Figure A.3: Map of counties included in matched analysis.** With the exception of California, which includes many primarily online IHEs, there are very few regions where the counties are clustered based on campus reopening strategy.

policies once they are in place. For instance, even in zip codes in states with mask mandates, we do not see full—or consistent—compliance from county to county [16]; similarly, we see differences in social distancing and mobility patterns among counties in the same state [17].

#### A.4 Campus COVID Dataset: Examples of IHE data

The Campus COVID Dataset includes detailed *time series* data about the number of tests and cases reported by the almost 1,000 IHEs. In Figures A.5, A.6, and A.7, we show three examples of IHEs that report daily case counts and tests conducted on campus. The shape of these three IHEs' testing curves exemplifies three broad trends seen in the testing cadence across IHEs during the Fall 2020 semester: For example, Northeastern University (Figure A.5) conducts a large number of tests throughout the semester, a rate that remains relatively high throughout September, October, and November. North Carolina State University at Raleigh (Figure A.6) reported thousands of "entry tests" early in the semester, followed by a relatively lower volume for the rest of the semester. University of California, Los Angeles (Figure A.7) dramatically increased its testing volume during late November, which followed

| variable | transformation | description | source |
|---|---|---|---|
| *County demographic variables* | | | |
| population density | log-scale | number of people per mile$^2$ | [9] |
| median income | log-scale | median income of residents in the county | [9] |
| population over 60 | per 100,000 county population; log-scale | sum of population in the following age brackets: 60-64, 65-69, 70-74, 75-79, 80-84, 85+ | [9] |
| urban/rural code | code = 4, 5, 6 $\rightarrow$ 1; code = 1, 2, 3 $\rightarrow$ 0 | Urban-Rural Classification (1: large central metro; 2: large fringe metro; 3: medium metro; 4: small metro; 5: micropolitan; 6: non-core). | [4] |
| *County COVID-19 response* | | | |
| stringency index (OxCGRT) | none | aggregate indicator of active mitigation policies, see Hale et al., (2021) | [8] |
| new COVID-19 tests | 7-day rolling avg.; per 100,000 county population; log-scale | daily number of COVID-19 diagnostic tests reported in the county | [10] |
| *IHE variables* | | | |
| fulltime enrollment | per 100,000 county population; log-scale | number of fulltime students enrolled at colleges in our dataset | [11] |
| fulltime enrollment (online) | per 100,000 county population; log-scale | number of fulltime students enrolled at colleges classified as "primarily online" | [5, 11] |
| fulltime enrollment (in person) | per 100,000 county population; log-scale | number of fulltime students enrolled at colleges classified as "primarily in-person" | [5, 11] |
| new COVID-19 tests | 7-day rolling avg.; per 100,000 county population; log-scale | sum of daily number of COVID-19 diagnostic tests reported by IHEs within the county | this work |
| *Other* | | | |
| 2020 voting behavior (% Rep.) | $\frac{\text{votes}_{Rep.}}{\text{votes}_{total}} \times 100$ | percent of total votes cast for the Republican candidate in the 2020 presidential election | [12] |
| average temperature (°Celsius) | none | daily average temperature, from the National Oceanic and Atmospheric Administration | [13] |
| *Dependent variable* | | | |
| new deaths from COVID-19 | 7-day rolling avg.; per 100,000 county population | daily number of reported deaths from COVID-19 at the county level | [10] |

**Table A.3: Description of variables in Table A.4.** Where appropriate, we use the "per 100k" designation—the variable's value divided by county population, multiplied by 100,000. Here "log" refers to the natural log, which we apply to variables that follow heavy-tailed distributions (e.g. income and population density).

a gradually increasing amount of tests reported throughout the semester. In each of the bar plots in this section, darker color bars indicate accelerating cases/tests.

Another common trend among the IHEs that conduct and report testing is an increase in testing volume prior to the Thanksgiving holidays (late November), which is when students

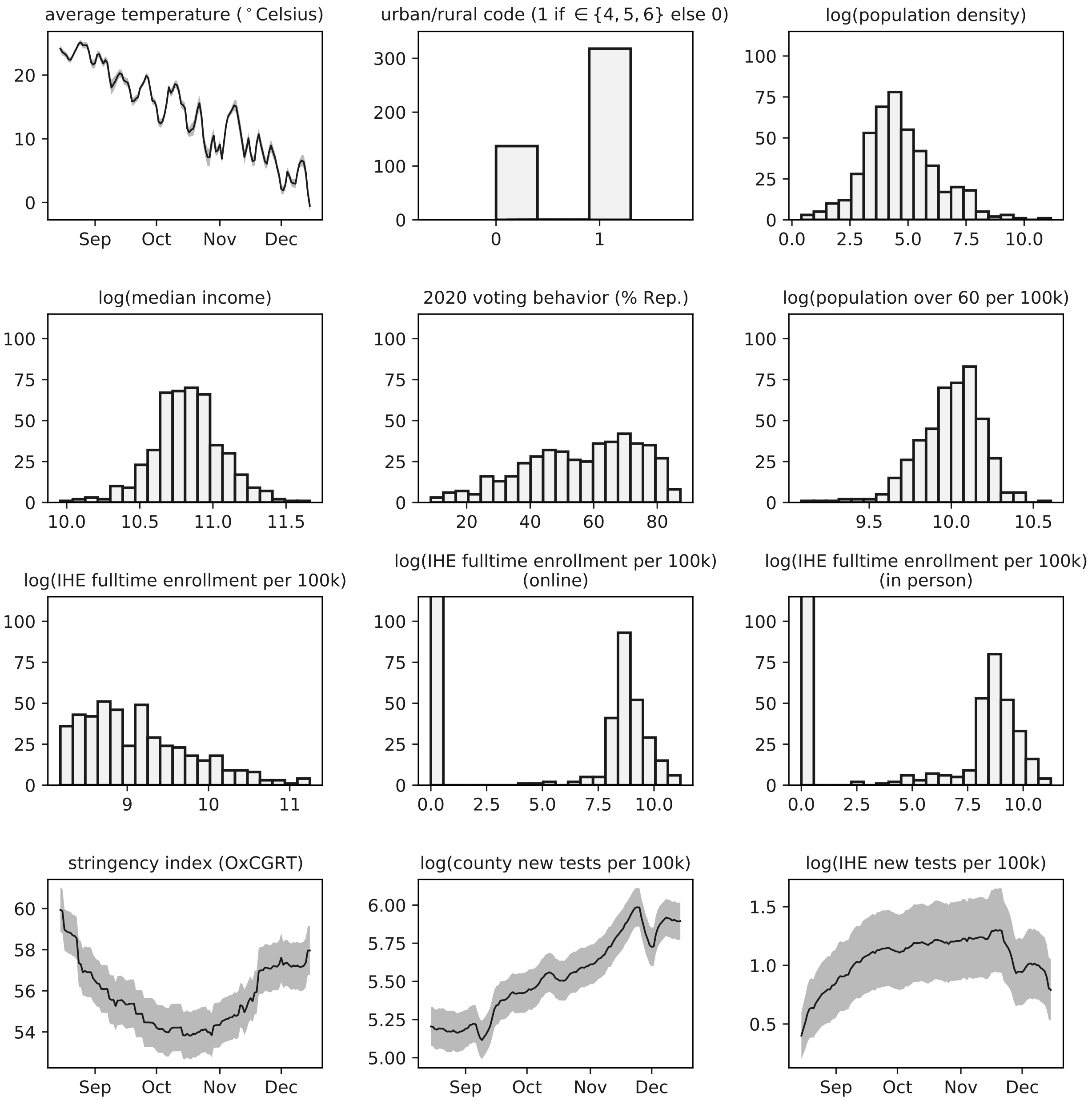

**Figure A.4: Distributions of the variables used in the regression in Table A.4.**

often return to their homes for several days. For Purdue University, University of Miami, and Georgia Institute of Technology (Figures A.8, A.9, and A.10), we see examples of these this mid-November spike in testing, followed by a sharp decline. This pattern is common across many IHEs, which often requested that students not return following this holiday, as the semester was already almost over and infection rates were beginning to surge in many parts of the United States.

Many IHEs only report cases and test counts in weekly intervals (e.g. Duke University

| | | | |
|---|---|---|---|
| **Dep. Variable:** | new deaths per 100k, at $t+38$ days | **No. Observations:** | 55842 |
| **Model:** | GLM | **Df Residuals:** | 55830 |
| **Model Family:** | NegativeBinomial | **Df Model:** | 11 |
| **Link Function:** | log | **Scale:** | 1.0000 |
| **Method:** | IRLS | **Log-Likelihood:** | -62440. |
| **No. Iterations:** | 8 | **Deviance:** | 31331. |
| **Covariance Type** | cluster (county) | **Pearson chi2:** | 4.09e+04 |

| | **coef** | **std err** | **z** | **P> \|z\|** | **[0.025** | **0.975]** |
|---|---|---|---|---|---|---|
| **average temperature (°Celsius)** | **-0.0391** | 0.003 | -14.917 | 0.000 | -0.044 | -0.034 |
| **urban/rural code (1 if $\in \{4,5,6\}$ else 0)** | 0.1014 | 0.070 | 1.442 | 0.149 | -0.036 | 0.239 |
| **log(population density)** | -0.0168 | 0.033 | -0.513 | 0.608 | -0.081 | 0.047 |
| **log(median income)** | **-0.4722** | 0.121 | -3.908 | 0.000 | -0.709 | -0.235 |
| **2020 voting behavior (% two-party vote)** | **0.0124** | 0.003 | 3.860 | 0.000 | 0.006 | 0.019 |
| **log(population over 60 per 100k)** | **0.4896** | 0.121 | 4.042 | 0.000 | 0.252 | 0.727 |
| **log(IHE fulltime enrollment per 100k)** | **-0.1028** | 0.049 | -2.084 | 0.037 | -0.199 | -0.006 |
| **log(IHE fulltime enrollment per 100k (online))** | 0.0088 | 0.010 | 0.890 | 0.374 | -0.011 | 0.028 |
| **log(IHE fulltime enrollment per 100k (in person))** | 0.0120 | 0.009 | 1.283 | 0.199 | -0.006 | 0.030 |
| **stringency index (OxCGRT)** | **-0.0178** | 0.004 | -4.171 | 0.000 | -0.026 | -0.009 |
| **log(county new tests per 100k)** | **0.2663** | 0.046 | 5.823 | 0.000 | 0.177 | 0.356 |
| **log(IHE new tests per 100k)** | **-0.0382** | 0.015 | -2.558 | 0.011 | -0.068 | -0.009 |

**Table A.4: Generalized Linear Model (GLM) Regression: Predicting COVID-19 deaths, at $t+38$ days.** Regression table under a Negative Binomial model. See Table A.3 & Figure A.4 for descriptions of variables. Standard errors were adjusted for clustering at the county level. Coefficients in **bold** are statistically significant.

and Ohio State University, Figures A.11 and A.12), and for hundreds of IHEs, we were only able to collect a summary of the cumulative number of tests conducted and cases reported throughout the semester.

These examples are not meant to be exhaustive—indeed many IHEs in the Campus COVID Dataset do not have time series data for their testing volume or case counts. It is our hope that by releasing these data and analyses that they will motivate other large scale data collection/standardization efforts moving forward. To view examples of time series from other IHEs or to download the entire Campus COVID Dataset, see [18].

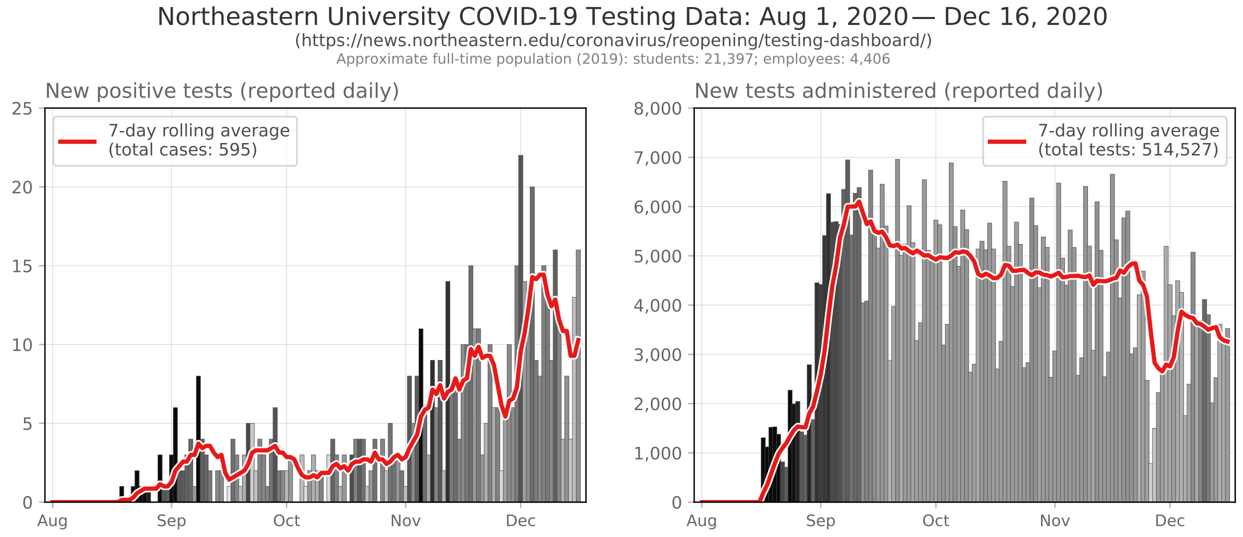

**Figure A.5: Example data: Northeastern University.**

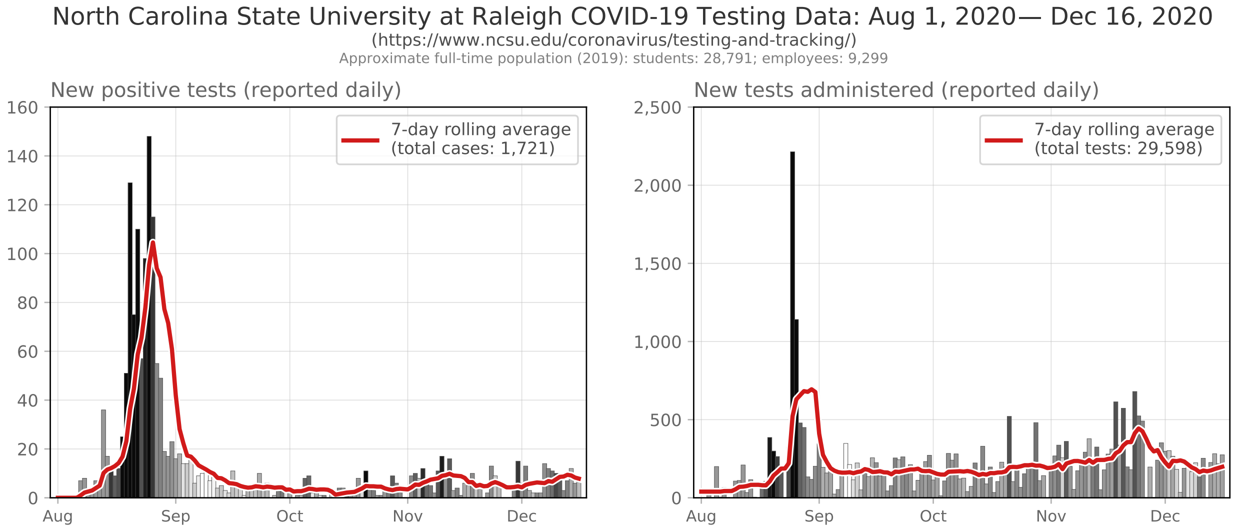

**Figure A.6: Example data: North Carolina State University at Raleigh.**

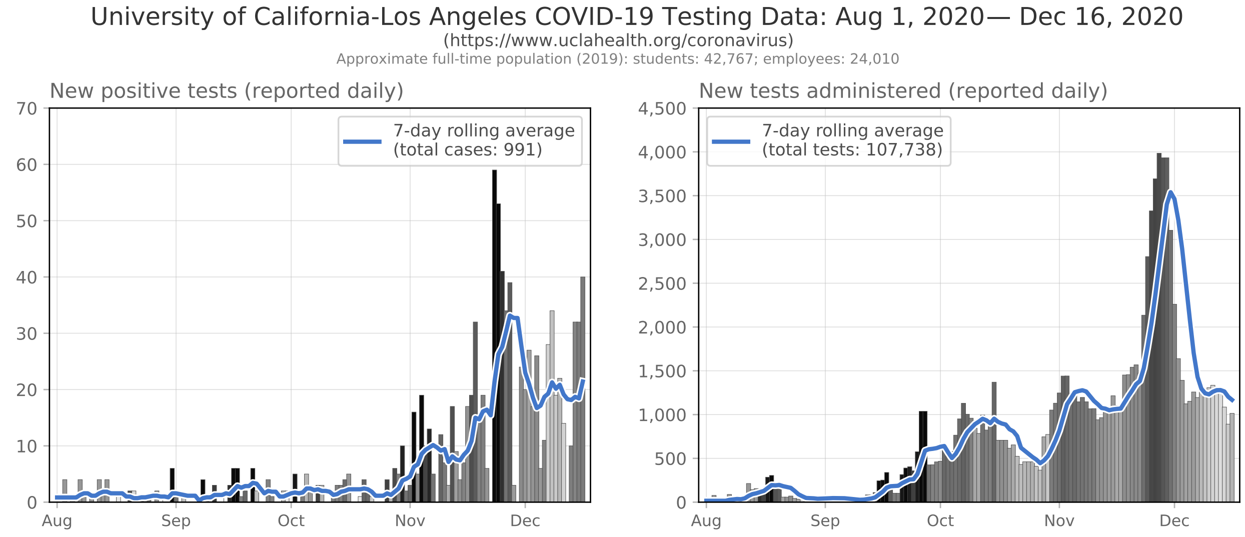

**Figure A.7: Example data: University of California-Los Angeles.**

[17] Brennan Klein, Tim LaRock, Stefan McCabe, Leo Torres, Lisa Friedland, Maciej Kos, Filippo Privitera, Brennan Lake, Moritz U.G. Kraemer, John S. Brownstein, David Lazer, Tina Eliassi-Rad, Samuel V. Scarpino, Alessandro Vespignani, and Matteo Chinazzi. "Quantifying collective physical distancing during the COVID-19 outbreak". In: *preprint* (2020). URL: https://www.mobs-lab.org/uploads/6/7/8/7/6787877/covid19mobility_report2.pdf.

[18] Brennan Klein. *jkbren/campus-covid: campus-covid*. Version v1.0. 2021. DOI: 10.5281/zenodo.5395340.

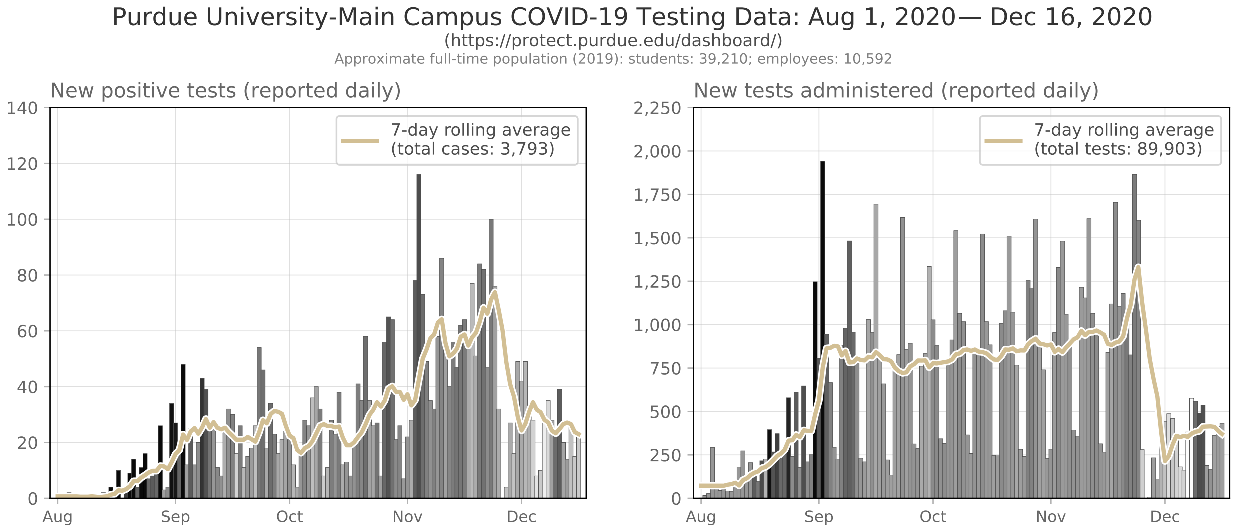

**Figure A.8: Example data: Purdue University-Main Campus.**

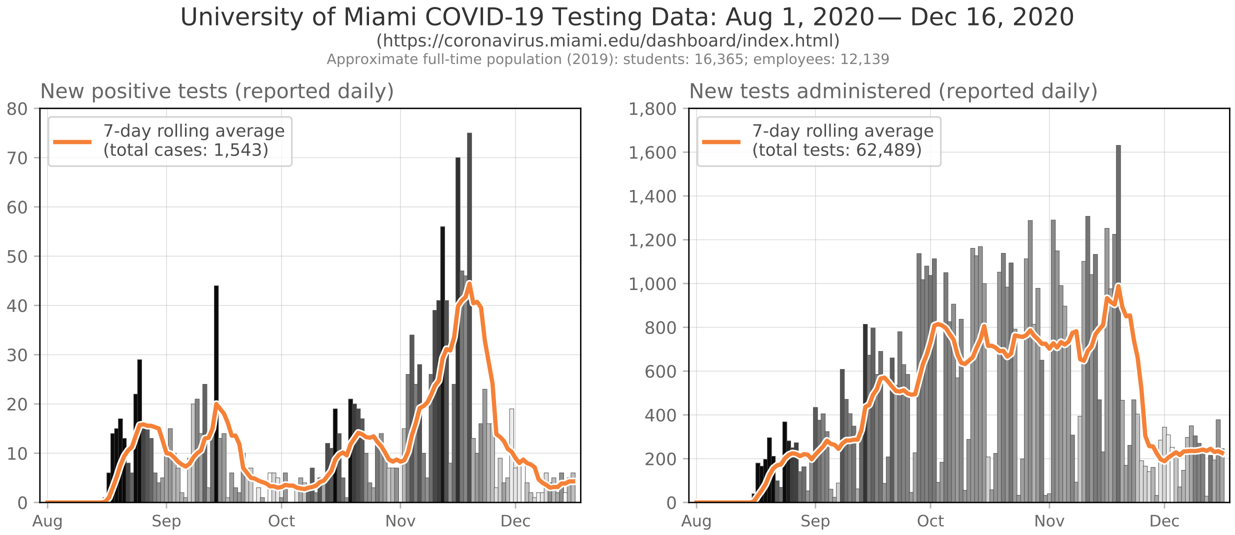

**Figure A.9: Example data: University of Miami.**

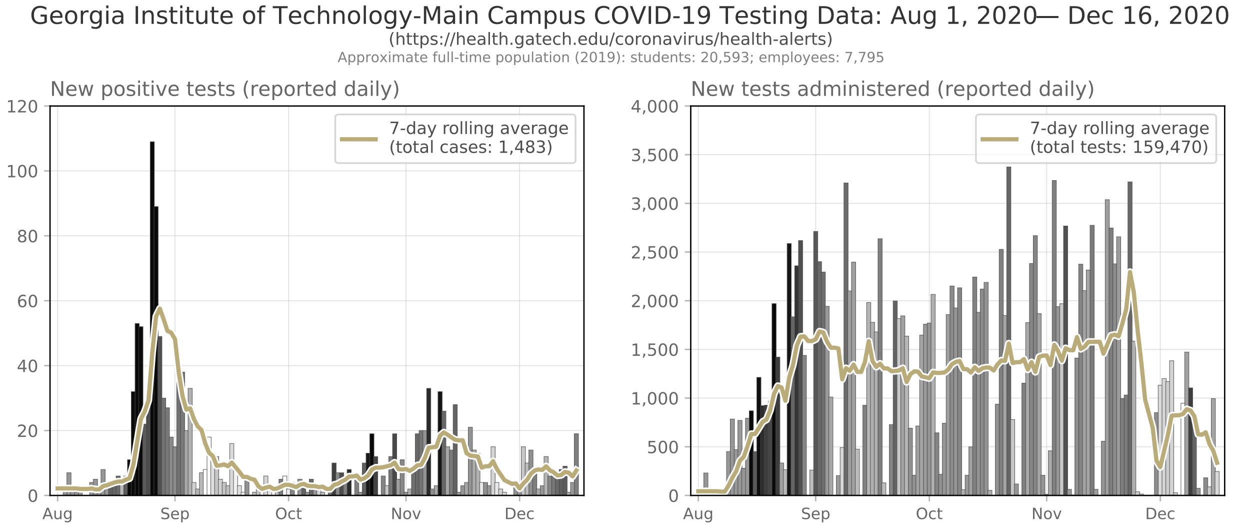

**Figure A.10: Example data: Georgia Institute of Technology-Main Campus.**

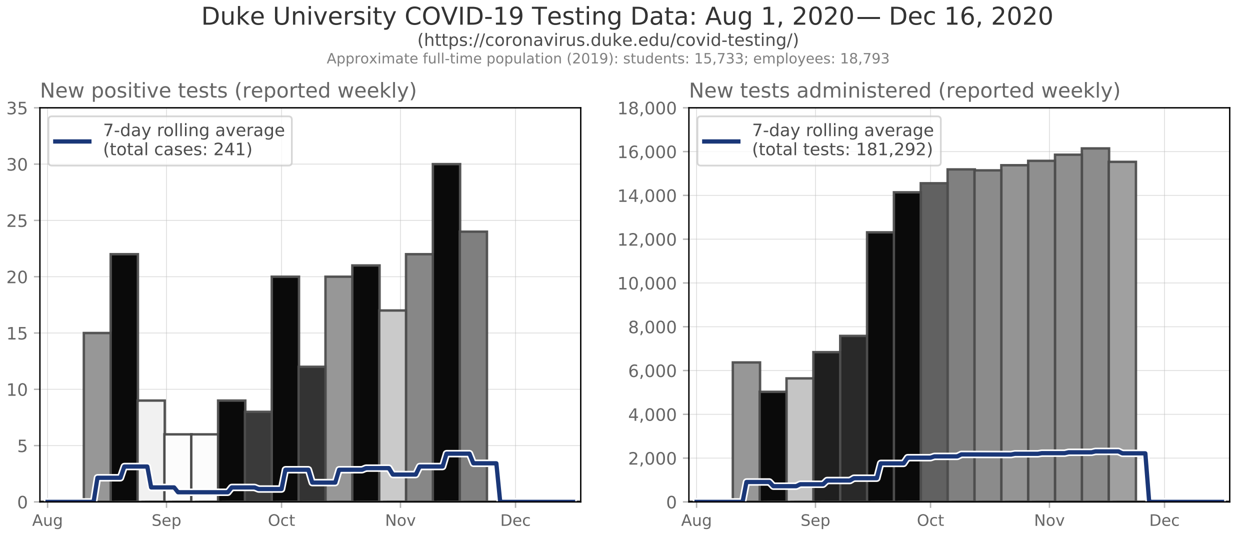

**Figure A.11: Example data: Duke University.**

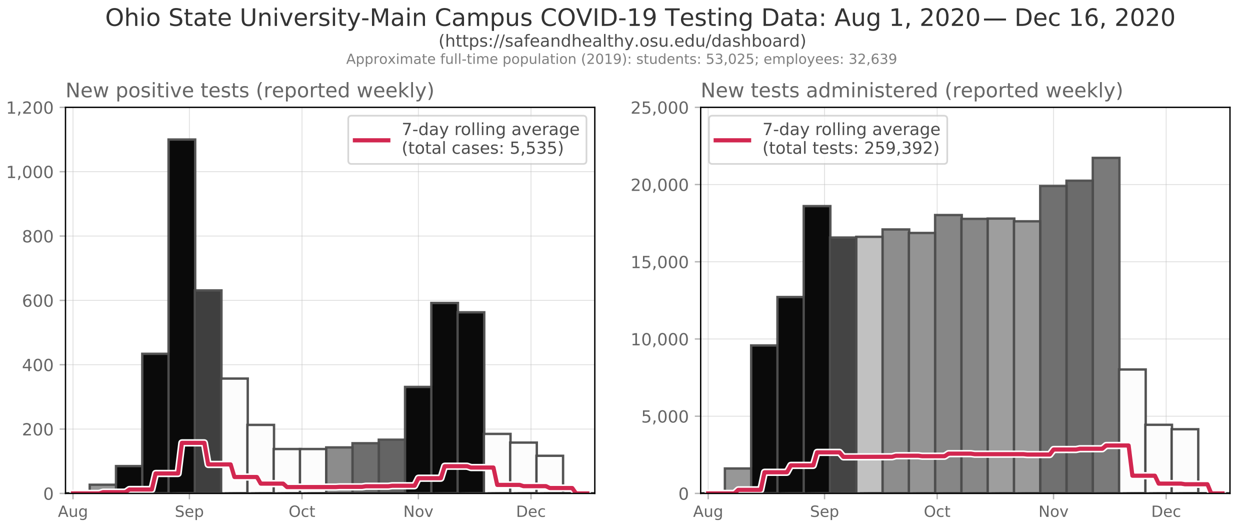

**Figure A.12: Example data: Ohio State University.**
